# Natural Language Processing to Build a Multicenter Computable Phenotype Library for Adults with Congenital Heart Disease

**DOI:** 10.1101/2025.08.18.25331953

**Authors:** Spencer Thomas, Angus Dawson, Hifsa Chaudhry, Sidra Ahmad, Xiyu Ding, Sarah A. Hummel, David M. Leone, Angela J. Weingarten, Eric Farber-Eger, Lauren Lee Shaffer, Benjamin P. Frischhertz, Sydney St. Clemmons, Sunil J. Ghelani, Fernando Baraona Reyes, Tzu-Chun Wu, Danny T. Y. Wu, Alexander R. Opotowsky, Timothy A. Miller

**Affiliations:** Computational Health Informatics Program, Boston Children’s Hospital, Boston, MA; Department of Cardiology, Boston Children’s Hospital, Boston, MA; Biomedical Informatics and Data Science (BIDS), Division of General Internal Medicine, Johns Hopkins University School of Medicine, Baltimore, Maryland, USA; Heart Institute, Department of Pediatrics, Cincinnati Children’s Hospital, University of Cincinnati College of Medicine, Cincinnati, OH; Vanderbilt University Medical Center, Nashville, TN; Harvard Medical School, Boston, MA; Department of Biostatistics, Health Informatics, and Data Sciences, University of Cincinnati, Cincinnati, OH; School of Information and Library Science (SILS), University of North Carolina, Chapel Hill, NC

**Author notes:** Correspondence to: Timothy Miller, Computational Health Informatics Program, Boston Children’s Hospital Department of Pediatrics, Department of Biomedical Informatics, Harvard Medical School, 401 Park Drive, Landmark Center, 5th Floor East, Boston, MA 02115, U.S.A. or Alexander R. Opotowsky, Cincinnati Adult Congenital Heart Disease Program, Heart Institute, Cincinnati Children’s Hospital, 3333 Burnet Avenue, MLC 2003, Cincinnati, OH 45229. ARO and TAM are co-senior authors.

**Keywords:** Adult congenital heart disease, natural language processing, machine learning, phenotyping

## Abstract

**Objective:** Our objective was to build classifiers for multiple phenotypes that categorize a cohort of adults with congenital heart disease (ACHD), that can be used to populate variables in a biobank.

**Materials and methods:** A dataset of 1492 ACHD patients, with expert-created labels for eight phenotypes, was created and used to train classifiers with three different architectures. A larger unlabeled dataset containing 15869 patients was used to pre-train the classifiers, and a 20% subset of the unlabeled dataset was used to validate the classifier predictions.

**Results:** On held out labeled data, F1 scores for the eight target phenotypes of interest ranged from 0.66 to 1. Of those, the six phenotypes with best classification performance were then validated on unlabeled data, where positive predictive value ranged from 81.5% to 100%.

**Discussion:** We were able to classify six out of eight phenotypes with satisfactory performance. Challenging phenotypes included cyanosis and New York Heart Association functional class. Both vary over time and in the latter case there is limited agreement between human observers. Different phenotypes benefited from different model architectures to some degree, but the differences are small enough that uniformity of deployment may be a more important factor in choosing what models to deploy. We saw no benefit to joint training, but some phenotypes benefited from a multiclass model.

**Conclusion:** Human-curated data can be used to train NTLP-based ACHD phenotype classifiers with excellent test characteristics acceptable for application in quality improvement efforts and to populate ACHD registry data.

## Introduction and background

Congenital heart disease (CHD) is defined as structural heart disease present at birth. These defects, present in about 1% of liveborn infants,[1] frequently require surgical or transcatheter interventions to allow survival. Historically, CHD has been considered a predominantly pediatric disease. Because of improved pediatric outcomes, however, there are now more adults than children alive with CHD. In 2010, there was an estimated 2.4 million individuals with CHD in the United States, and of these, nearly 59% were adults.[2] These adults are aging and surviving longer than ever. Given their unique characteristics, differences of their disease relative to acquired cardiovascular disease, and the ongoing growth of this population, there is a significant gap in understanding their medical needs.

### Objective

The objective of this study is to create algorithmically-derived labels for eight high-yield phenotypes of interest not dependably identified using structured medical record data in a cohort of ACHD patients. We take advantage of unique manually curated data resources at a tertiary care pediatric hospital, along with recent methods in natural language processing, to create phenotype classifiers for our eight expert-derived categories and test classifier architectures with a range of complexity to find the best-performing models. The models we train can be deployed as software containers with simple interfaces and are being deployed at validation sites to contribute to a multi-site ACHD database.

## Methods and materials

Our patient population is derived from a large tertiary care pediatric hospital. We make use of two research databases: The first dataset is a **labeled** cohort of 1492 patients enrolled in an ongoing ACHD Biobank.[3] These individuals had unstructured clinical notes available in a research data warehouse along with detailed forms completed by congenital cardiologists to characterize their cardiac diagnoses. The second dataset is **unlabeled** and includes 15869 patients who were seen at any time in the ACHD program, meet predefined coded criteria (described below), and have unstructured clinical notes stored in the same research data warehouse. See **Figure 1** for a flowchart describing the cohort.

**Figure 1:**
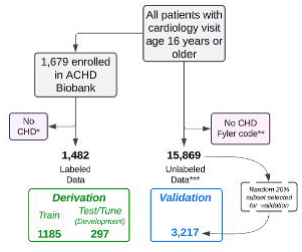
Flowchart of cohort for derivation and validation of adult congenital heart disease computable phenotypes. * *Examples: Marfan syndrome, pulmonary hypertension without CHD. ** See Supplemental Table 1. *** Labeled only with data available in the medical record, including extensive CHD labeling with Boston Children’s Hospital Fyler codes.* This flowchart describes the cohort used for training, testing/tuning (development), and internal validation of computable phenotypes.

Our dataset is unique in that it contains coded data for each encounter in the form of ***Fyler codes***. Fyler codes are a cardiology-specific set of codes that was developed at Boston Children’s Hospital and that are still in use.[4] While these codes are mapped to standard SNOMED codes for billing purposes, they contain more granular detail than typical administrative data and contain further clinical details. These Fyler codes were used to extract the **unlabeled** dataset, with the specific codes (and their descriptions) included in supplementary materials.

### Labeled dataset

The labeled dataset was created as part of the Boston ACHD Biobank, which enrolled outpatients with CHD aged ≥18 years between 2012 and 2019 seen at Boston Children’s or Brigham and Women’s Hospitals; a detailed description of the protocol has been published elsewhere. Written informed consent was obtained from each participant. The study was approved by Boston Children’s Hospital’s Institutional Review Board, and there was a formal reliance agreement with the Partners HealthCare/Brigham and Women’s Hospital Institutional Review Board.

This project used a REDCap database[5] including demographic and clinical data manually collected from electronic medical records via detailed review of problem lists, medication lists, procedural and imaging reports, and available clinical notes. Data were collected around the time of enrollment, reflecting the clinical status on the date of baseline biospecimen collection. In the absence of a relevant cardiovascular intervention (e.g., valve replacement), data obtained within 2 years of sample collection were included. Extensive clinical data was extracted from the medical record, with >826 questions/fields providing detailed descriptions of the patient’s cardiac history and current status. Information included CHD diagnoses, demographics, anthropometric data, vital signs, New York Heart Association (NYHA) functional class, laboratory results, cardiac imaging results, and cardiopulmonary exercise testing results.[3] Data for each patient was extracted/annotated by two separate individuals (research coordinators or investigators), and discrepancies were adjudicated by a third individual. We selected eight initial phenotypes to develop classifiers using variables in our labeled REDCap database, based on clinician input for impact and feasibility. The chosen phenotypes were pulmonary hypertension (PH), cyanosis, New York Heart Association functional class (NYHA) functional class, congenitally corrected transposition of the great arteries (cc-TGA), dextro-transposition of the great arteries (D-TGA), Fontan circulation (Fontan), Eisenmenger syndrome, and a history of atrial arrhythmia (AA).

These terms were defined using the criteria described in Table 1. D-TGA and cc-TGA are mutually exclusive in our labeling schema (i.e., a patient can have one or neither, but never both), and there is nothing to be gained by modeling them separately, so we combined them into one TGA classifier to avoid possible errors where patients were predicted as having both diagnoses.

**Table 1:**
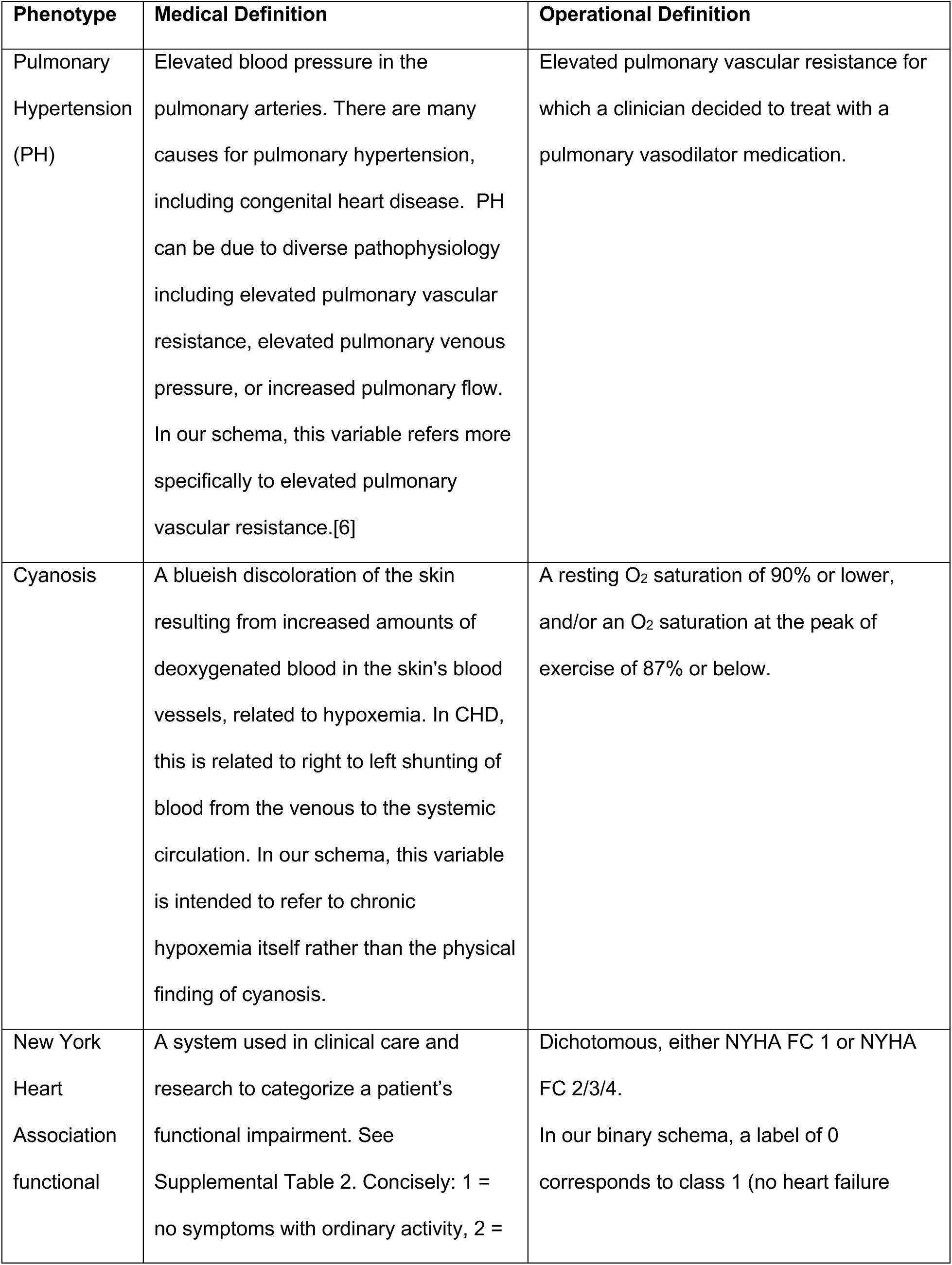
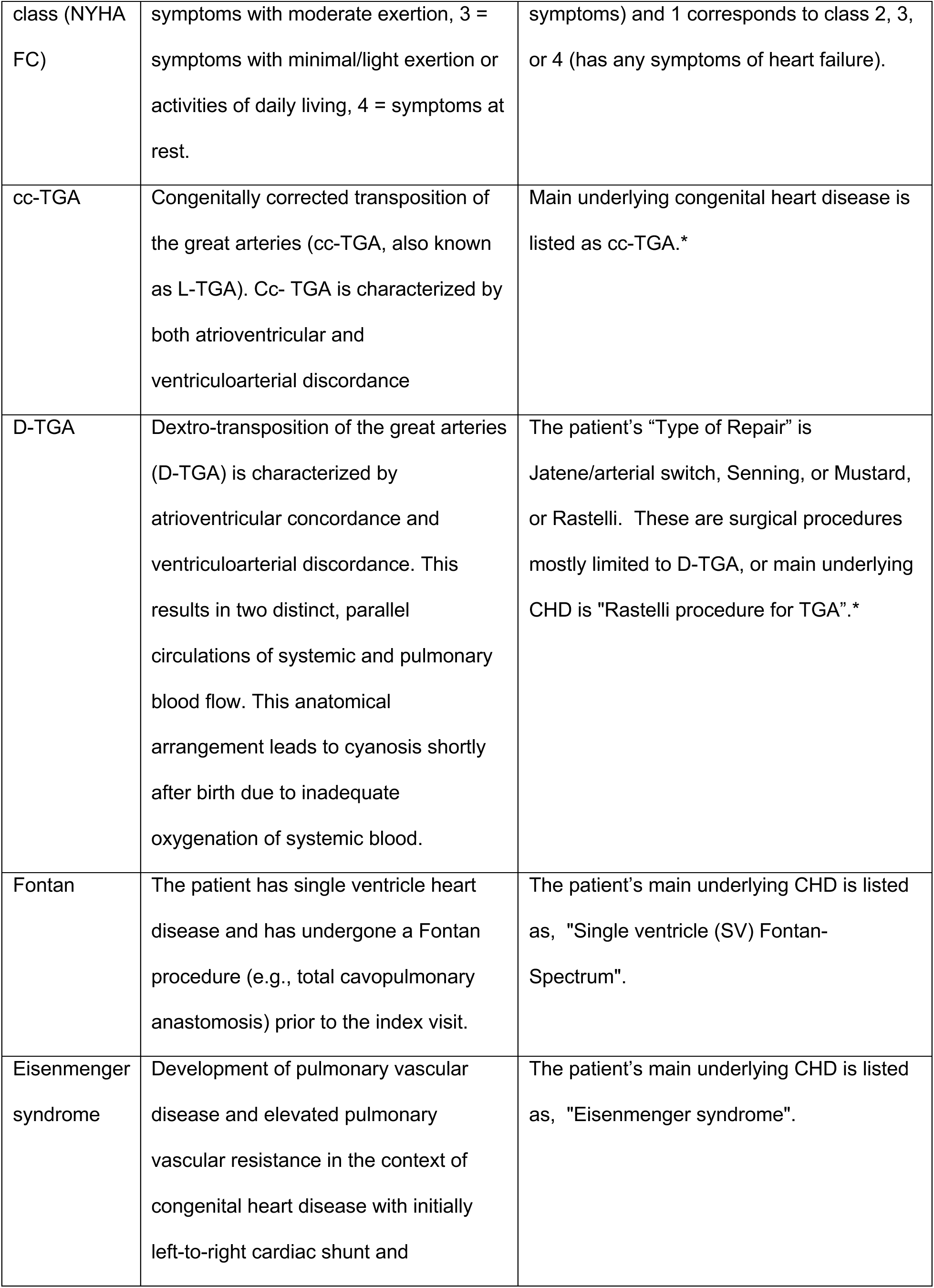
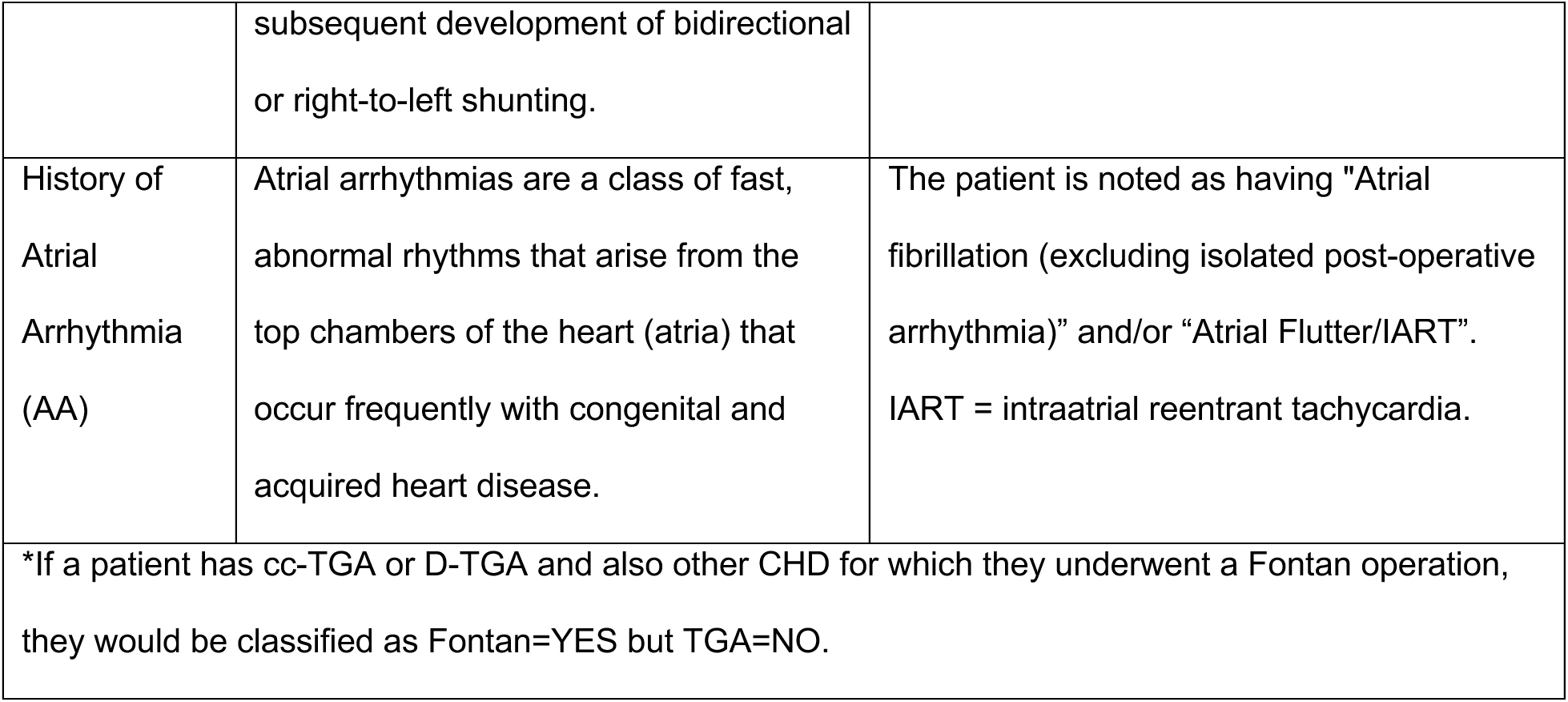
Phenotype labels, medical definitions, and operational definitions.

We separated the labeled data into training and development splits for developing machine learning models. Because some phenotypes had relatively few positive cases, separate training and development sets were created for each phenotype, stratified relative to that label, to ensure an adequate number of positive examples were included in both the training and development sets.

### Unlabeled dataset

We also constructed an unlabeled dataset intended to represent the target population for future classification using validated phenotype classifiers. To maximize sensitivity, patients were selected using broad inclusion criteria, accepting some degree of false positives. Specifically, we selected all patients who were seen in clinic at age ≥16 years between 1990 and 2020 and matched a broad set of Fyler codes that could potentially be indicative of any type of congenital heart disease (See supplementary materials for a full list). For this set of patients, we queried research databases for all encounters, all accompanying free text notes, labs, procedures, demographic data, and codes (both Fyler and International Classification of Diseases).

### Classifier development

Classifier development proceeded in a simple-to-complex fashion, using methods that were suited for long document classification. The first baseline we employed uses a bag-of-words feature extractor and a support vector machine (SVM) classifier with linear kernel. The next classifier we explored was a convolutional neural network [7] with a randomly initialized embedding layer. Finally, we explored a hierarchical version of a Bidirectional Encoder Representations from Transformers (BERT) classifier.[8] A more traditional BERT classifier could not be applied to this data because our dataset contained very long documents that do not fit into the context window.

#### Support Vector Machine (SVM)

TF-IDF is a simple text embedding scheme composed of a document’s term frequency (TF), the number of times a given token appears in the document, divided by inverse document frequency (IDF), the number of documents in which that term appears. TF-IDF representations for each text were fed into a linear model with stochastic gradient descent training. A hyperparameter grid-search was conducted to find the optimal regularization coefficient, loss penalty, and class weighting scheme, in addition to whether to exclude stop words. The optimal hyperparameter settings can be found in the supplementary materials.

#### Convolutional Neural Network (CNN)

A hyperparameter grid-search was conducted to find the optimal embedding size, filter sizes, and number of filter layers for each phenotype. After the best-performing model from the above experiments was selected, another hyperparameter grid search was conducted for each phenotype to find the optimal batch size and learning rate. Learning rate scheduling was employed such that learning rate decreased throughout training.

Most high-performing models had between 500 and 1000 filter layers, an embedding size of 50 or 100, a base learning rate of 0.005-0.01, and a batch size of 16. The optimal hyperparameter sets can be found in the supplementary materials.

#### Hierarchical Clinical NLP Transformer (HierCNLPT)

This final classifier used a pre-trained BERT encoder to encode chunks of 512 tokens, and has a transformer at a higher layer that takes as input the encoded first token of each chunk.[9]

Since the higher-level transformer is randomly initialized, we pre-trained the hierarchical transformer model using Fyler codes as our “ground truth” for training. To build this pre-training dataset, we used the unlabeled dataset to create a set of labeled instances, with an output space of size 1000 sigmoid classifiers, representing the 1000 most frequently used Fyler codes.

During pre-training, the classifier is trained end-to-end so that the chunk encoder weights (i.e., the weights of the pre-trained BERT encoder) can also be updated during training. We explored treating this model as a feature extractor, where the network weights would be frozen, and patient vectors could be extracted by passing in text.

However, this approach showed poor performance in preliminary work. We therefore focused on a fine-tuning approach in which a classifier layer specific to each phenotype was added to the document-level transformer’s final layer, using the first classifier chunk as its input representation. This allowed for end-to-end fine-tuning, resulting in distinct trained models for each phenotype (HierCNLPT+FT).

#### Validation

A subset of the unlabeled dataset, containing 3217 patients, was processed by the best-performing CNN models for the different pre-selected phenotypes: AA, Fontan, Eisenmenger, TGA, and PH. Each individual model generated a list of predicted positive patients. These phenotypes were validated by clinical research assistants with a background in biology and trained by a board-certified cardiologist available for periodic review and consultation regarding difficult to adjudicate cases. From this verification, we were able to determine the positive predictive value for each of these models.

## Results

Table 1 shows demographic information of the labeled dataset and unlabeled dataset, overall and the subset used for initial validation, as well as cohort size and note lengths.

**Table 1:** Demographics (Data are shown as median [1st, 3rd quartile] or n (%).

| **Table 1: Demographics (Data are shown as median [1st, 3rd quartile] or n (%)).** |  |  |  |
| --- | --- | --- | --- |
|  | **Labeled data** | **Unlabeled data** | **Unlabeled (subset)** |
| Age at baseline encounter (years) | 41 [33, 55] | 34 [26, 45] | 34 [26,44] |
| Race, *n* (%) | 1492 (100%) | 15869 (100%) | 3217 (100%) |
| White | 1067 (71.51%) | 9811 (61.82%) | 2010 (62.48%) |
| Hispanic | 63 (4.22%) | 980 (6.18%) | 190 (5.91%) |
| Black | 30 (2.01%) | 734 (4.63%) | 135 (4.20%) |
| Asian | 25 (1.68%) | 341 (2.15%) | 75 (2.33%) |
| Multiracial | 4 (0.27%) | 67 (0.42%) | 14 (0.44%) |
| Other | 37 (2.48%) | 512 (3.23%) | 111 (3.45%) |
| Unknown/Missing | 266 (17.83%) | 3424 (21.57%) | 682 (21.20%) |
| Sex, *n* (%) |  |  |  |
| Female | 758 (50.80%) | 7974 (50.25%) | 1615 (50.20%) |
| Male | 733 (49.13%) | 7894 (49.75%) | 1602 (49.80%) |
| Unknown/ Missing | 1 (0.07%) | 1 (0%) | 0 (0%) |
| **Note length (tokens)** | 10471 [4076,<br>22573] | 5707 [1778,<br>20526] | 5795 [1818, 19768] |

### Labeled dataset

Tables 2 and 3 show the class distributions for the seven phenotypes of interest. The least skewed label distribution is binary NYHA functional class, where 21.44% of the training set and 28.09% of the development set are in the positive class.

**Table 2:** Class representations for binary phenotypes. AA = atrial arrhythmias, NYHA FC = New York Heart Association functional class, PH= pulmonary hypertension.

| Phenotype, *n* (%) | Train |  | Dev |  |
| --- | --- | --- | --- | --- |
|  | Negative | Positive | Negative | Positive |
| AA | 956 (80.13%) | 237 (19.87%) | 240 (80.27%) | 59 (19.73%) |
| Cyanosis | 1032 (94.33%) | 62 (5.67%) | 259 (94.53%) | 15 (5.47%) |
| Eisenmenger | 1184 (99.25%) | 9 (0.75%) | 297 (99.33%) | 2 (0.67%) |
| Fontan | 1034 (86.67%) | 159 (13.33%) | 259 (86.62%) | 40 (13.33%) |
| NYHA FC > 1 | 894 (78.56%) | 244 (21.44%) | 215 (71.91%) | 84 (28.09%) |
| PH | 1157 (96.98%) | 36 (3.02%) | 290 (96.99%) | 9 (3.01%) |

**Table 3:**
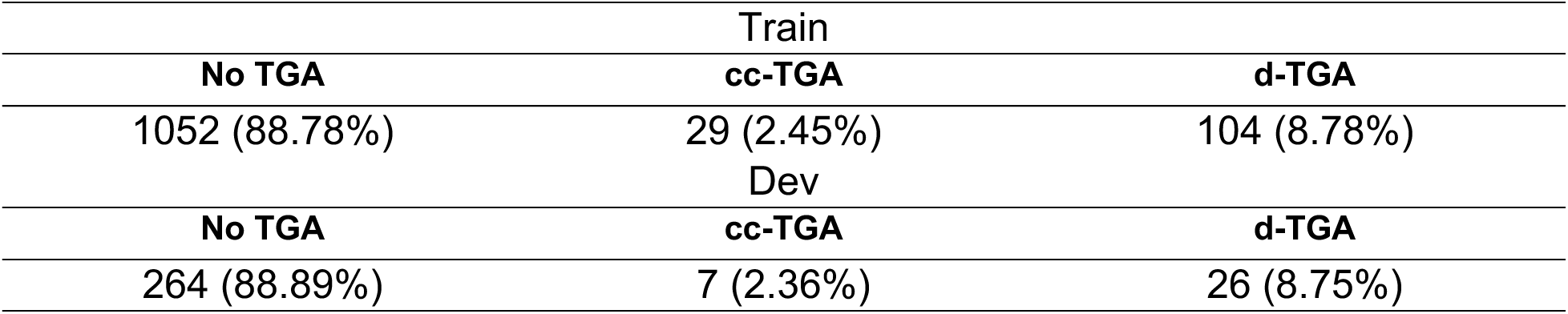
Class representations for the TGA phenotypes (TGA=Transposition of the great arteries, cc-TGA=congenitally corrected TGA, d-TGA=dextro-TGA).

Table 4 shows the positive class F1 in the best-performing model in each category. The best model is that which achieves the highest F1 in the positive class of the held-out development dataset. Precision and recalls for this classifier are in Supplemental Table 2.

**Table 4.**
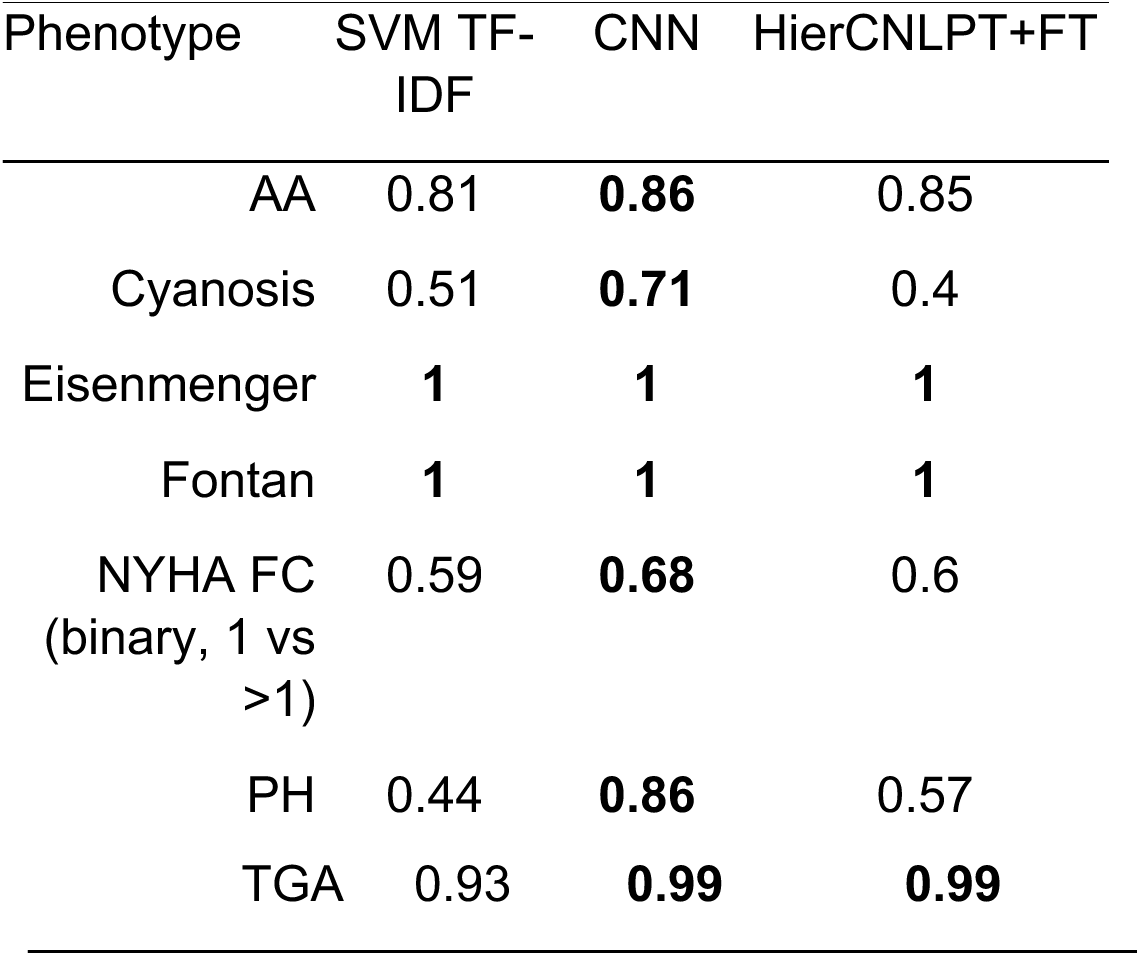
F1 results of 3 different model architectures. AA = atrial arrhythmias, NYHA FC = New York Heart Associated functional class, PH = pulmonary hypertension, TGA=Transposition of the great arteries.

Table 5 shows the results of validation of the classifier predictions on a subset of the unlabeled data. Because of the large number of negative cases, it was not possible to validate the sensitivity of the classifiers.

**Table 5.** Validation results on a portion of the unlabeled dataset. PPV=Positive predictive value. PPV on dev set is included as a point of comparison to the labeled dataset. Cc-TGA and d-TGA were validated separately as binary predictions despite the original classifier being multi-class.

| Phenotype | # Predicted cases | # True positives | PPV | PPV on dev set |
| --- | --- | --- | --- | --- |
| AA | 162 | 132 | 81.48% | 87.72% |
| cc-TGA | 30 | 26 | 86.67% | 100% |
| d-TGA | 150 | 128 | 85.33% | 100% |
| Eisenmenger | 8 | 8 | 100% | 100% |
| Fontan | 160 | 154 | 96.25% | 100% |
| Pulmonary hypertension | 19 | 17 | 89.47% | 100% |

The CNN architecture showed the most promising results on the labeled development set, while also offering substantially lower computational costs; therefore, it was selected for the validation experiments. However, it is noteworthy that the HierCNLPT model achieved near-perfect performance on TGA, and all three architectures performed perfectly on Fontan and Eisenmenger phenotypes.

In the validation experiments, all classifiers had PPV greater than 80%, with Eisenmenger and Fontan classifiers nearly perfect. PPV on the validation set was in the same range as PPV on the development set. For the Fontan validation, we note that 9 patients had a heart transplant after their Fontan procedure, and 4 had a Fontan takedown (i.e., the patients no longer have Fontan physiology). Both categories were treated as true positives, as the model correctly recognized that the patients had had a Fontan procedure, even if they no longer have Fontan physiology.

## Discussion

We were able to complete our goal to create machine-learning derived natural language processing classifiers to identify eight complex, and distinct ACHD phenotypes. Of these, 6/8 reached our pre-specified benchmark performance metrics. Fontan circulation had the best success with a near perfect ability to distinguish the condition.

In contrast, classification of NYHA functional class proved substantially more difficult, with performance not exceeding an F1 score of 0.66; even when reframed as a binary task (Class I vs. Class II/III/IV) Early experiments explored joint training of all classification models, based on the hypothesis that a single model performing multiple phenotype classification tasks could benefit from shared representation learning in the lower layers of the network. However, this approach showed no performance advantage. We did observe modest benefit from training a multiclass model specifically for TGA phenotypes, by merging the classifier’s upper decision-making layers rather than its lower feature-extraction layers. Because D-TGA and cc-TGA labels were mutually exclusive in the dataset, this setup forced the model to assign a single label, thereby reducing false positives in which a patient was simultaneously flagged for both subtypes—a limitation encountered when using separate binary classifiers. This strategy is not universally applicable, but it suggests there may still be performance gains available with a structured output space.

The two classifiers with the worst performance, cyanosis and NYHA FC, provide a greater challenge for binary classification. Both vary substantially over time and can be subjective by the observer. For cyanosis, individual patients often had multiple oxygen saturation measurements on the same day or during the same visit, with some values meeting criteria for cyanosis and others not. In addition, provider documentation may reflect varying thresholds for defining cyanosis in clinical notes, introducing further inconsistency in the labeling. The binary classifier limits classification accuracy in such a context. Future efforts should include integration of variation over time and possibly a third level for identifying patients with variable or borderline cyanosis or NYHA FC. The limited accuracy of the NYHA FC classifier is further explained by the intrinsic subjectivity of classification and well-described poor inter-observer agreement in assigning NYHA FC.[10]

This work explored a range of simple to complex classifiers for each phenotype. There have been focused efforts to develop NLP-based classifiers for related fields,[11] and for an individual ACHD phenotype,[12] but to our knowledge no similar work developing multiple classifiers in parallel. Our original intention was to use the simplest classifier for each phenotype that performs sufficiently well. However, there is a competing interest of simplicity of deployment that suggests using the single architecture that performs best on average. This may lead to some (simpler) phenotypes having more complex classifiers than necessary, and some (more challenging) phenotypes sacrificing performance for uniformity of deployment.

These classifiers perform well and, pending external validation, appear sufficiently accurate for many real-world applications. However, the inherent complexity of clinical phenotype definitions necessitates human review in certain contexts, depending on the intended use. For example, our model labeled any individual with a history of Fontan circulation as a positive case, regardless of subsequent heart transplant or Fontan takedown. This approach is appropriate for research or quality improvement efforts focused on Fontan outcomes, but such patients should be identified and excluded when defining a population for active management within a Fontan follow-up program.

One limitation is that we could not realistically validate the full set of test characteristics in the unlabeled validation cohort (n=3217, a 20% sample of the total unlabeled dataset). The proportion of positive cases was substantially lower in the validation data for each phenotype. For example, Fontan comprised 13.33% of the labeled dataset vs. 4.79% of unlabeled cohort (based on identified true positive cases). The labeled dataset was not a random sample of the overall population, however; this cohort would be expected to be enriched for the phenotypes of interest for several reasons including that they tend to be more complex and would be more likely to be enrolled in a prospective study since they remain in specialized care with frequent clinical follow-up. This could potentially lead to unrealistically high estimates of case counts if the classifier overfit to the prevalence in the labeled data. However, our validation results show high PPVs, ranging from 81.48% to 100%, suggesting that our classifiers were able to generalize to a dataset with a different label distribution.

The differences between labeled and unlabeled datasets are also apparent in the substantially longer notes in the former. Inclusion in the labeled dataset was generally predicated on enrollment in ACHD clinic, where notes tend to be much longer than in pediatric cardiology clinics. The labeled dataset is also, relative to the unlabeled dataset, likely to under-represent patients with limited available information (e.g., a small number of visits, only distant visits), those seen in the pediatric cardiology clinic only, and those with mild disease. These differences provided an opportunity to validate the algorithms in a population meaningfully different than the derivation cohort; taken in that context, the favorable test characteristics of the algorithms in the unlabeled dataset are especially reassuring.

### Conclusion

We were able to develop six high performing natural language processing-based classifiers for ACHD phenotypes. More complex methods offer occasional benefits, but these benefits may not be worth the complexity of training and deployment. Future directions are aimed at validation and improving model performance.

## Supporting information

Supplemental data file with Fyler codes

## Data Availability

The primary data produced in the present study is not able to be made publicly available, because it contains patient details that cannot be legally or ethically shared.

## Abbreviations

AA: history of atrial arrhythmia
ACHD: adult congenital heart disease
BERT: Bidirectional Encoder Representations from Transformers
CNN: Convolutional neural network
HierCNLPT+FT: Hierarchical clinical natural language transformers model with fine tuning
CHD: congenital heart disease
PH: pulmonary hypertension
NYHA FC: New York Heart Association functional class
TGA: Transposition of the great arteries
cc-TGA: congenitally corrected transposition of the great arteries
d-TGA: dextro-TGA (complete transposition of the great arteries)
MRN: medical record number
SVM TF-IDF: Support vector machine classifier with term frequency-inverse document frequency-based features

