## Supplemental Table 1 and 2 for "Natural Language Processing to Build a Multicenter Computable Phenotype Library for Adults with Congenital Heart Disease"

Supplemental materials:

**S.1: Hyper-parameter tuning details**

**Supplemental Table 1**. Best-performing hyperparameters. For Fontan and Eisenmenger CNN classifiers, multiple hyperparameter configurations yielded the best result; we have included the lowest such values.

SVM

| Phenotype | Alpha | Penalty | Class weight | Exclude stopwords |
| --- | --- | --- | --- | --- |
| AA | 0.001 | L1 | balanced | True |
| Cyanosis | 0.01 | L2 | balanced | True |
| Eisenmenger | 0.01 | L1 | balanced | True |
| Fontan | 0.0001 | L1 | balanced | False |
| NYHA FC | 0.001 | L1 | balanced | False |
| PH_1 | 0.01 | L2 | balanced | True |
| PH_2 | 0.001 | L2 | balanced | False |
| TGA | 0.0001 | L1 | None | True |

CNN

| Phenotype | Embedding size | # of filters | Filter sizes | Learning rate | Batch size |
| --- | --- | --- | --- | --- | --- |
| AA | 100 | 250 | 2,3,4 | 0.01 | 16 |
| Cyanosis | 100 | 1000 | 1,3,5 | 0.01 | 16 |
| Eisenmenger | 25 | 250 | 1,3,5 | 0.01 | 16 |
| Fontan | 25 | 250 | 1,3,5 | 0.005 | 8 |
| NYHA FC | 50 | 250 | 2,3,4 | 0.005 | 32 |
| PH1 | 25 | 1000 | 2,3,4 | 0.01 | 16 |
| PH2 | 25 | 500 | 1,3,5 | 0.1 | 16 |
| TGA | 100 | 100 | 2,3,4 | 0.01 | 16 |

HierCNLPT

For the HierCNLPT experiments, we searched over learning rate, and all models performed the best on the development set with a learning rate of 5e-5.

**Supplemental Table 2**: Precision and recall values (dev set)

| Phenotype | Minority class recall | Minority class precision |
| --- | --- | --- |
| AA | .88 | .85 |
| Cyanosis | .77 | .67 |
| Eisenmenger | 1 | 1 |
| Fontan | 1 | 1 |
| NYHA_FC | .6 | .79 |
| PH | 1 | .75 |
| TGA | 1 | .98 |

**S.2: Fyler codes used to extract unlabeled data:**

See attached spreadsheet for a description of all Fyler codes, and whether they were used as inclusion criteria to define the unlabeled cohort.
